# PACT-3D, a Deep Learning Algorithm for Pneumoperitoneum Detection in Abdominal CT Scans

**DOI:** 10.1101/2024.03.01.24303638

**Authors:** I-Min Chiu, Teng-Yi Huang, Kuei-Hong Kuo

## Abstract

Pneumoperitoneum, necessitates surgical intervention in 85-90% of cases, relies heavily on CT scans for diagnosis. Delay or misdiagnosis in detecting pneumoperitoneum can significantly increase mortality and morbidity. Our study introduced PACT-3D, a deep learning model developed to identify pneumoperitoneum in CT images. In this single hospital study, we retrospectively reviewed abdominal CT scans from January 2012 to December 2021, excluded CT of image acquisition error and without reports to form the development dataset for training the model. We evaluated the PACT- 3D model using a simulated test set of 14,039 scans and a prospective test set of 6,351 scans, collected from December 2022 to May 2023 at the same center. PACT-3D achieved a sensitivity of 0.81 and a specificity of 0.99 in retrospective testing, with prospective validation yielding a sensitivity of 0.83 and a specificity of 0.99. Sensitivity improved to 0.95 and 0.98 when excluding cases with a small amount of free air (total volume < 10ml) in simulated and prospective test sets, respectively. By delivering accurate and consistent patient-level predictions and providing segmented masks, PACT- 3D holds significant potential for assisting rapid decision-making in emergency care, thereby potentially improving patient outcomes.

## Introduction

Pneumoperitoneum, which refers to the presence of extraluminal free air in the peritoneal space, is a potentially life-threatening condition that represents a differential diagnosis when managing acute abdominal pain in the Emergency Department (ED). In adults, perforated viscus is the leading cause of pneumoperitoneum, representing 85-95% of cases, and among these, surgical pneumoperitoneum comprises 85-90%^1,2^. Diagnostic tools for identifying pneumoperitoneum include plain radiographs, ultrasound, and Computed Tomography (CT) scan, with the latter remaining the gold standard, exhibiting reported sensitivity levels of approximately 96-100%^3^. Timely diagnosis of pneumoperitoneum is crucial, as delayed recognition can lead to sepsis and result in increased mortality and morbidity^4,5^. However, prolonged CT interpretation times are frequently observed in crowded EDs, with previous reports indicating an average delay of approximately 2 hours^6^. Moreover, the use of CT scans during ED visits has dramatically increased in the past decade, with a 330% rise reported in the US from 3.2% of encounters (95% confidence interval [CI] 2.9% to 3.6%) in 1996 to 13.9%^7^.

Diagnosing pneumoperitoneum from a CT scan is highly dependent on the reader’s expertise and the amount of free air present. According to previous research, only 62.8% of junior physicians feel confident about diagnosing acute pathological findings from CT scans, such as pneumoperitoneum or bowel obstruction^8^. Moreover, studies have shown that discrepancy rates in the interpretation of emergency CT scans between residents and attending radiologists vary significantly based on the level of training, ranging from 13.5% to 30.0%^9,10^. Misinterpretations can have a direct negative impact on patient management, with adverse effects noted in 7.2% of patients^11^. These factors may contribute to considerable delays in the recognition of critical pathologies like pneumoperitoneum, potentially leading to poorer patient outcomes.

Artificial Intelligence (AI) has greatly advanced healthcare in recent years, particularly in medical imaging technologies such as computed tomography (CT) scans, X-rays, and ultrasonography^12-16^. AI has also contributed to increased speed and efficiency in medical image analysis, reducing the workload of healthcare professionals and improving patient outcomes. Recent studies have investigated the potential of deep learning algorithms in assisting the detection of pneumoperitoneum on CT scans^17,18^. However, the performance of these AI models varies and is dependent on the selection of datasets. For assessment of the AI model, it is critical to use a dataset that mirrors the actual incidence rate of pneumoperitoneum. Moreover, a prospective evaluation is necessary, along with ongoing enhancements to improve model performance.

In this study, we introduced PACT-3D, a 3-dimensional U-Net algorithm is a convolutional neural network architecture specifically tailored for 3D medical image segmentation, excelling in capturing spatial hierarchy and information across both the transverse and vertical axes of biomedical images. The PACT-3D model is designed to automatically segment areas of pneumoperitoneum from CT scans, providing predictions at the patient level and visualizations at the pixel level. It is engineered to detect pneumoperitoneum with high accuracy, and its performance has been thoroughly evaluated using both a simulated test dataset and in a prospective observational setting.

## Result

### 3.1 Demographic Characteristics

In this study, we retrospectively analyzed 140,339 abdominal CT scans from 2012 to 2021. After exclusions, 139,781 were eligible for analysis. Pneumoperitoneum was identified in 973 of these and the studies were randomly allocated to training, validation, and test datasets in an 8:1:1 ratio (Figure 1). The training set comprised 695 scans with pneumoperitoneum, alongside a randomly selected equivalent number of negative scans. The validation set included 139 scans with pneumoperitoneum, matched with an equal number of negative cases. To evaluate the performance of the PACT-3D model, the test set was designed to mirror a real-world prevalence ratio of approximately 1:100, consisting of 139 scans with pneumoperitoneum and a larger pool of 13,900 negative scans. Additionally, we conducted a prospective clinical evaluation using abdominal CT scans from December 2022 to May 2023 at the same hospital, resulting in a prospective test set of 6,351 CT scans. This approach aims to thoroughly evaluate the model’s performance under conditions that closely resemble those of clinical settings.

**Figure 1.**
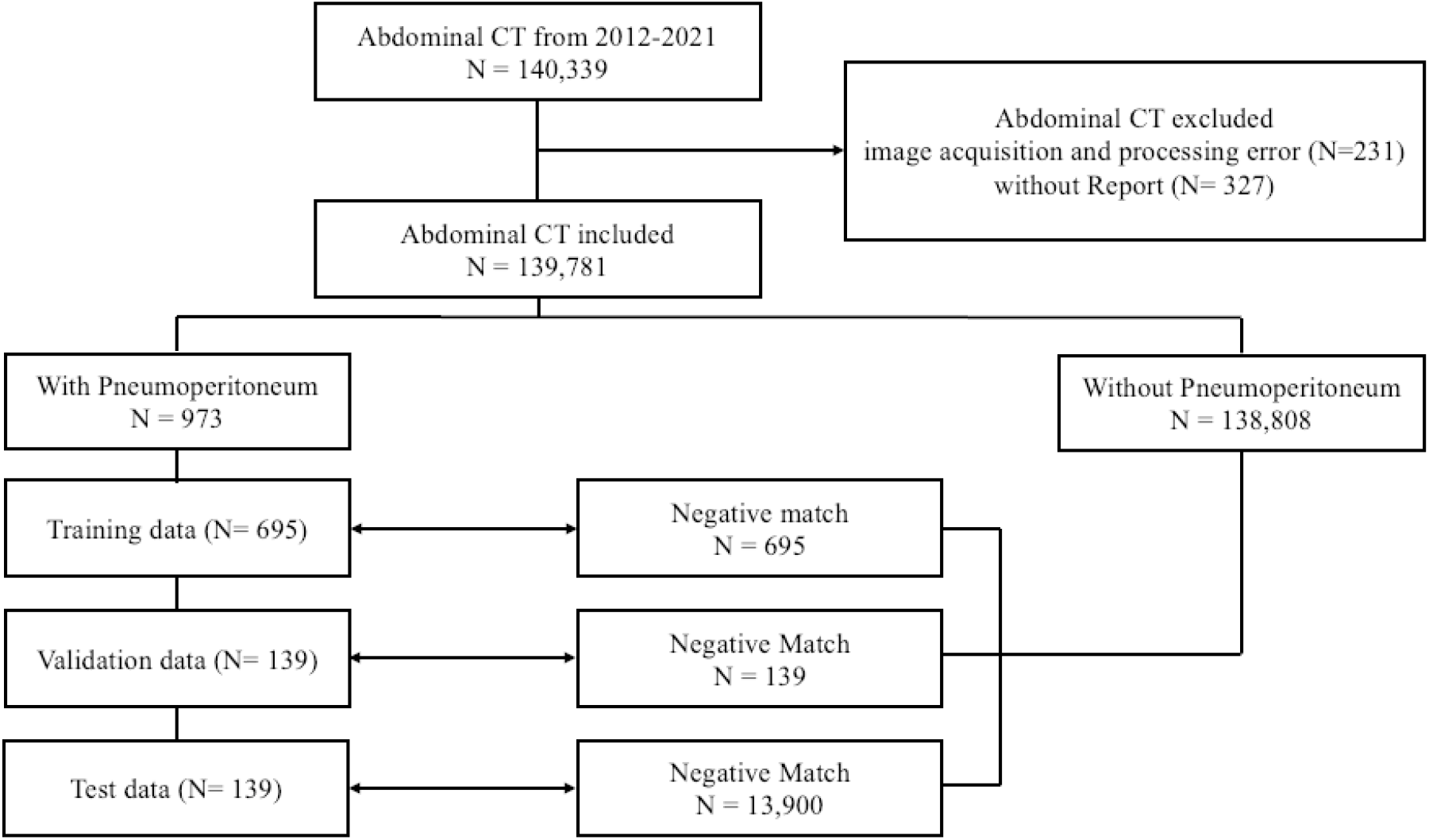
The inclusion flowchart of this study.

The mean age of patients in the simulated test set was 54 years with a standard deviation (SD) of 13.1, while the prospective test set had a slightly higher mean age of 59 years (SD = 16.9). Females represented 48.2% (n=6,767) of the simulated test set and 47.2% (n=3,000) of the prospective test set. The incidence of pneumoperitoneum detected was set to 1.0% in the simulated test set. Analyzing all CT scans in ER, the incidence of pneumoperitoneum was 1.3% (n=82) in the prospective test set (Table 1).

**Table 1.** Demographics and CT Vendor distributions in Simulated and Prospective Test Sets.

|  | **Simulated Test Set**<br>Mean (SD) / N (%) | **Prospective Test Set**<br>Mean (SD) / N (%) |
| --- | --- | --- |
| **Total CT scans** | 14,039 | 6,351 |
| **Age** | 54 (13.1) | 59 (16.9) |
| **Female** | 6,767 (48.2%) | 3,000 (47.2%) |
| **CT Vendors** |  |  |
| Philips Brilliance 64 | 1,123 (8.0%) |  |
| Siemens Somatom<br>definition | 1,502 (10.7%) |  |
| Siemens Somatom<br>definition Flash | 772 (5.5%) | 524 (8.3%) |
| Siemens Somatom<br>definition AS | 624 (60.1%) | 2,772 (43.6%) |
| GE LightSpeed VCT | 2,204 (15.7%) | 1,479 (23.3%) |
| GE Revolution<br>Frontier |  | 1,576 (24.8%) |
| **Pneumoperitoneum** | 139 (1.0%) | 82 (1.3%) |

CT=Computed Tomography; PPV=Positive Predictive Value

### 3.2 Distribution of CT Vendors

Regarding the distribution of CT vendors, there were noticeable differences between the simulated and prospective test sets. In the simulated test set, Philips Brilliance 64 scanners were used in 8.0% of cases, while Siemens Somatom Definition and Definition Flash scanners were used in 10.7% and 5.5% of cases, respectively. GE LightSpeed VCT scanners accounted for 15.7% of the scans. A significant portion, 60.1%, involved Siemens Somatom Definition AS scanners (Table 1).

In contrast, the prospective test set exhibited a varied distribution. Siemens Somatom Definition AS scanners were used less frequently, constituting 43.6.9% of the scans. The GE Revolution Frontier became more prevalent, representing 24.8% of scans in this set. This shift in vendor distribution indicates a temporal change in scanner preference or availability between the two test sets. The image acquisition setting of different CT vendors was shown in Supplemental Table 1.

### 3.3 Model Performance

The trained 3D U-Net model demonstrated satisfactory performance in detecting pneumoperitoneum on the validation set. The Dice score for pneumoperitoneum segmentation was 0.81, indicating a high degree of overlap between the predicted and ground truth regions. Throughout the training process, we meticulously balanced the number of negative CT scans against positive ones at varying ratios to refine the model’s sensitivity and positive predictive value (PPV). Our objective was to optimize the F1-score, which harmonizes sensitivity and PPV, as reflected in Supplemental Table 2. The data revealed that a balanced ratio of positive to negative cases (1:1) yielded the highest F1-score.

In the simulated test set, our model achieved a F1-score of 0.54 (95% CI: 0.47-0.61), with a sensitivity of 0.81 (95% CI: 0.75-0.86), a specificity of 0.99 (95% CI: 0.98-1.0), and a PPV of 0.41 (95% CI: 0.34-0.38). Of the 139 CT scans positive for pneumoperitoneum, the model identified 112 and missed 27. Among the 13,900 negative scans, 167 were incorrectly classified as pneumoperitoneum. In the prospective test set at ER, the model’s performance yielded F1-score of 0.58 (95% CI: 0.51-0.65), with a sensitivity of 0.83 (95% CI: 0.77-0.90), specificity of 0.99 (95% CI: 0.98-0.99), and a PPV of 0.44 (95% CI: 0.37-0.52). Out of the 69 CT scans with confirmed pneumoperitoneum, the model detected 54 and misclassified 88 out of 8,451 negative scans (Table 2).

**Table 2.** Performance of PACT-3D in Test Set.

| Performance Metrics | Simulated Test Set | Prospective Test Set |
| --- | --- | --- |
|  | value (95% CI) | value (95% CI) |
| Sensitivity | 0.81 (0.75-0.86) | 0.83 (0.77-0.90) |
| Specificity | 0.99 (0.98-1.0) | 0.99 (0.98-0.99) |
| PPV | 0.41 (0.34-0.48) | 0.44 (0.37-0.52) |
| F1-score | 0.54 (0.47-0.61) | 0.58 (0.51-0.65) |
| Sensitivity in etiology |  |  |
| Gastro-duodenal | 0.93 (0.82-0.98) | 0.87 (0.73-0.94) |
| Small Bowel | 1.0 (0.87-1.0) | 0.88 (0.63-0.98) |
| Large Intestine | 0.64 (0.41-0.77) | 0.73 (0.50-0.89) |
| Trauma | 1.0 (0.57-1.0) | 0.83 (0.45-0.97) |
| Post-operative | 0.59 (0.33-0.84) | 0.8 (0.55-0.93) |
| Sensitivity in total volume of free air |  |  |
| Total volume > 1ml | 0.89 (0.84-0.93) | 0.91 (0.86-0.95) |
| Total volume > 10ml | 0.95 (0.90-0.98) | 0.98 (0.93-1.0) |

### 3.4 Subgroup analysis

When analyzing performance by etiological subgroup, the PACT-3D model displayed remarkable accuracy for gastroduodenal and small bowel perforations, as well as trauma-related cases, achieving sensitivities of 0.93 (0.82-0.98), 1.0 (0.87-1.0), and 1.0 (0.57-1.0), respectively. In contrast, the model demonstrated relatively lower sensitivities for large intestine perforation and post-operative changes, recording values of 0.64 (0.41-0.77), 0.59 (0.33-0.84). During the prospective observational period, a consistent pattern was observed. The sensitivities for gastroduodenal, small bowel, and trauma-related perforations were 0.87 (0.73-0.94), 0.88 (0.63-0.98), and 0.83 (0.45-0.97), while those for large intestine perforation and post-operative changes were 0.73 (0.50-0.89) and 0.8 (0.55-0.93), respectively (Table 2).

In subgroup analyses evaluating performance across various total volumes of free air, we observed improvement in the sensitivity of PACT-3D. Specifically, sensitivity increased to 0.89 (95% CI: 0.84-0.93) and 0.91 (95% CI: 0.86-0.95) on the simulated and prospective test sets, respectively, when scans with a total free air volume of less than 1ml were excluded. This sensitivity further escalated to 0.95 (95% CI: 0.90-0.98) and 0.98 (95% CI: 0.93-1.0) upon excluding scans with less than 10ml of total free air volume, indicating a correlation between detection capability and the quantity of free air present (Table 2).

From another point of view, an association was found between scans predicted as positive by the model and a heightened rate of urgent surgeries, defined as surgeries conducted within 24 hours following the CT scan. After excluding post operation scans, in the simulated test set, urgent surgeries were performed on 84 (85.8%) of the patients out of 99 whose pneumoperitoneum was identified by the model. In contrast, among the patients with missed pneumoperitoneum diagnoses by the model, 10 (55.6%) out of 18 underwent urgent surgeries (p<0.001). Within the prospective test set, 40 (75.5%) of the 53 patients diagnosed with pneumoperitoneum by the model received urgent surgeries, as opposed to 8 (57.1%) of the 14 patients with pneumoperitoneum that the model failed to detect (p<0.001).

## Discussion

In this study, we introduced PACT-3D, a 3D U-Net-based deep learning model, designed for detecting pneumoperitoneum on abdominal CT scans. The robustness of PACT-3D is underlined by its training on scans from a wide array of CT scanner models and its validation across different time periods, ensuring consistent performance despite the evolving landscape of medical imaging technology. PACT-3D demonstrated exemplary performance, characterized by high sensitivity and specificity. The model’s high specificity and satisfactory PPV are particularly noteworthy given the rarity of pneumoperitoneum in routine settings, which is crucial for minimizing false positives and thus reducing the risk of alarm fatigue. The consistent performance of PACT-3D, observed through its application on a prospective test set that included newer CT scanner models, further substantiates its reliability and potential for aiding clinical decision-making across various clinical scenarios and timeframes. Historically, AI algorithms have encountered challenges when attempting to detect free air in CT scans. They often exhibit reduced sensitivity, even if their specificity is commendable^17,18^. Previous studies, focusing on the utilization of 2D segmentation models for pneumoperitoneum detection, have highlighted challenges in differentiating free air from the common place bowel gas^18^. While 2D models have been a cornerstone in healthcare deep learning applications, this is largely because many medical imaging modalities, such as X-rays, ultrasound, and specific MRI or CT slices, intrinsically generate 2D images, making these models a natural choice^19-22^. Additionally, 2D models tend to be computationally less demanding than 3D models, suiting institutions with restricted computational capabilities. The extensive availability of pretrained 2D models, which have been trained on diverse and vast datasets, further contributes to their dominance^23^. By fine-tuning these models for specific medical tasks, performance can often be enhanced, benefiting from features learned across various domains. However, despite the prevalence of 2D architectures in healthcare, the detection of pneumoperitoneum, with its inherent risk of confusion with bowel air, greatly benefits from the depth of understanding offered by 3D morphology. Our employment of a 3D segmentation model permits superior recognition of free air morphological patterns, distinguishing them from bowel gas with enhanced accuracy. This adaptation, coupled with the model’s rapid inference capability, heightens its potential to augment diagnostic precision and efficiency.

In the subgroup analysis, PACT-3D particularly excelled in detecting gastroduodenal and small bowel origin pneumoperitoneum, with sensitivities exceeding 0.9 in both test sets. For large intestine origin cases, sensitivity ranged between 0.64 and 0.73. We surmise this disparity arises from the inherently larger air bubble sizes in the upper gastrointestinal tract, facilitating differentiation from standard bowel gas. In contrast, large intestine perforations, frequently linked with inflammatory processes, present greater interpretative challenges, even for seasoned radiologists^24,25^. Consistent with this, the model demonstrated improved sensitivity when CT scans with minimal free air volume were excluded, showing an increase to 0.89-0.91 for total free air volumes greater than 1ml, and further to 0.95-0.98 for volumes greater than 10ml (Table 2).

When assessing the model’s predictions in relation to clinical outcomes, specifically the necessity for urgent surgery, we observed a significant correlation. Patients with pneumoperitoneum detected by the PACT-3D model underwent urgent surgery at a higher rate (75.5-85.8%) compared to those where pneumoperitoneum was not detected (55.6- 57.1%). This suggests that the model is more adept at identifying larger volumes of free air, particularly those originating from the upper gastrointestinal tract, where emergency surgical intervention is often imperative. Conversely, smaller volumes of free air, typically resulting from inflammatory conditions like acute diverticulitis, are usually managed with conservative treatment or elective surgery in patients who are hemodynamically stable^26^. These findings indicate that the PACT-3D model can also serve as a valuable tool for risk stratification by illustrating the perforated area alongside the volume of free air.

Comparatively, our model’s performance aligns with or exceeds prior deep learning endeavors in detecting pneumoperitoneum or related abdominal pathologies on CT scans. This corroborates the robustness and superiority of our 3D U-Net-based approach. The remarkable sensitivity and specificity further posit PACT-3D as a potent ally for radiologists, especially under emergent circumstances where prompt and accurate detection is paramount. Several facets contribute to the success of PACT-3D, including the implementation of the 3D U-Net architecture, renowned for its efficacy in diverse medical image segmentation tasks, and the amalgamation of Dice loss and focal loss to counteract training set imbalances.

Several limitations are inherent to our study. Firstly, our research was conducted within the confines of a single medical center, potentially constraining the generalizability of our findings. While PACT-3D demonstrated proficiency within this setting, its applicability across varied clinical landscapes remains to be seen. It would be prudent for future endeavors to evaluate PACT-3D’s performance using datasets spanning multiple institutions, ensuring a more comprehensive representation of patient demographics and imaging variations. Secondly, while our model showcased robust performance in detecting pneumoperitoneum, the specific efficacy in discerning smaller or more subtle instances was not rigorously assessed. Given that such nuanced cases often present a diagnostic challenge, future research should zero in on the model’s prowess in these scenarios, determining its true potential as a diagnostic aid.

Despite the limitations of our study, the clinical implications of PACT-3D are profound. By serving as an auxiliary tool our model can potentially refine diagnostic accuracy, enhance radiologist or first line clinician’s confidence. In our future studies, the aim will be to validate the model across diverse populations and clinical settings, and to evaluate its contribution to improving clinical workflow.

In conclusion, this study highlighted the feasibility of developing a deep learning model that accurately identify pneumoperitoneum in abdominal CT scans. As a 3-dimensional model in medical image segmentation, PACT-3D maintained consistent performance across different testing periods. Its high specificity helps to avoid clinician fatigue due to false alarms, while its high sensitivity is particularly noteworthy in cases with larger volumes of free air. The model holds significant potential to aid rapid decision-making in emergency care, which could lead to improved patient outcomes.

## Method

### 2.1 Study Setting

In our research, we employed a dataset of post contrast abdominal CT scans from a single medical center, collected over a period spanning from January 2012 to December 2021. This dataset was enriched with CT scans indicating the presence or absence of pneumoperitoneum, a condition diagnosed using formal radiologist reports. For scans identified as positive for pneumoperitoneum, verification was performed by two radiologists who confirmed the presence of free air during the annotation process. To assess its applicability in a clinical setting, the model was prospectively validated from December 2022 to May 2023 in the same hospital for its performance in real-world data. This study was reported under the STARD protocol and received approval from the Institutional Review Board of Far Eastern Memorial Hospital (FEMH 111086-F). All participant records were de-identified and anonymized prior to analysis.

### 2.2 Image Data Acquisition

Abdominal CT scans during the study period were collected, and so does the corresponding reports. We included only the CT scans with contrast injection, axial plane scan, and reformatting slice thickness of 5 mm, with the field of view including the abdomen. CT scans with image acquisition and processing error, and CT scan without reports were excluded from this study. Figure 1 illustrates the recruitment and analysis flowchart.

### 2.3 Dataset Collection and Splitting

We employed natural language processing (NLP) methods to retrieve reports with and without a positive description of pneumoperitoneum from the image database. Initially, we utilized the NLP results as CT labeling and subsequently made minor revisions based on a random check of 1/5 of the CTs. We enrolled all CT scans that displayed pneumoperitoneum. The data was divided into training, validation, and test sets in a 5:1:1 ratio. To ensure no data leakage, CT scans from the same patient were exclusively allocated to the training set. For CT scans without pneumoperitoneum, we randomly selected non-duplicated patient scans, ensuring a 1:1 match with the pneumoperitoneum scans for both the training and validation sets. To mimic real-world conditions, our test set was formulated with a clinical ratio of 1:100 for positive to negative cases, reflecting an annual prevalence.

### 2.4 Image Annotation

Two senior radiologists with both 13 years of experience radiologist manually segmented the free gas bubble on the axial section with a window width and center of 600 HU and 40 HU, respectively. Contouring of bowel gas was strictly prohibited. Later, the labeled pixels with CT number of corresponding image higher than -150HU were removed. Finally, another radiologist checked and revised all pneumoperitoneum annotations. Prior to using the data for training, we standardized all CT images by removing the window width settings and applying pixel normalization based on the maximal and minimal values.

### 2.5 Deep Learning Model and Training

For pneumoperitoneum segmentation, we developed a 3D U-Net based neural network to predict the segmented mask of bowel gas (Figure 2)^27^. Its design incorporates a contracting path to capture context, juxtaposed with a symmetric expanding path, which facilitates precise localization. In enhancing the network, successive layers replace traditional pooling operations with up-sampling operators, thereby refining the output resolution.

**Figure 2.**
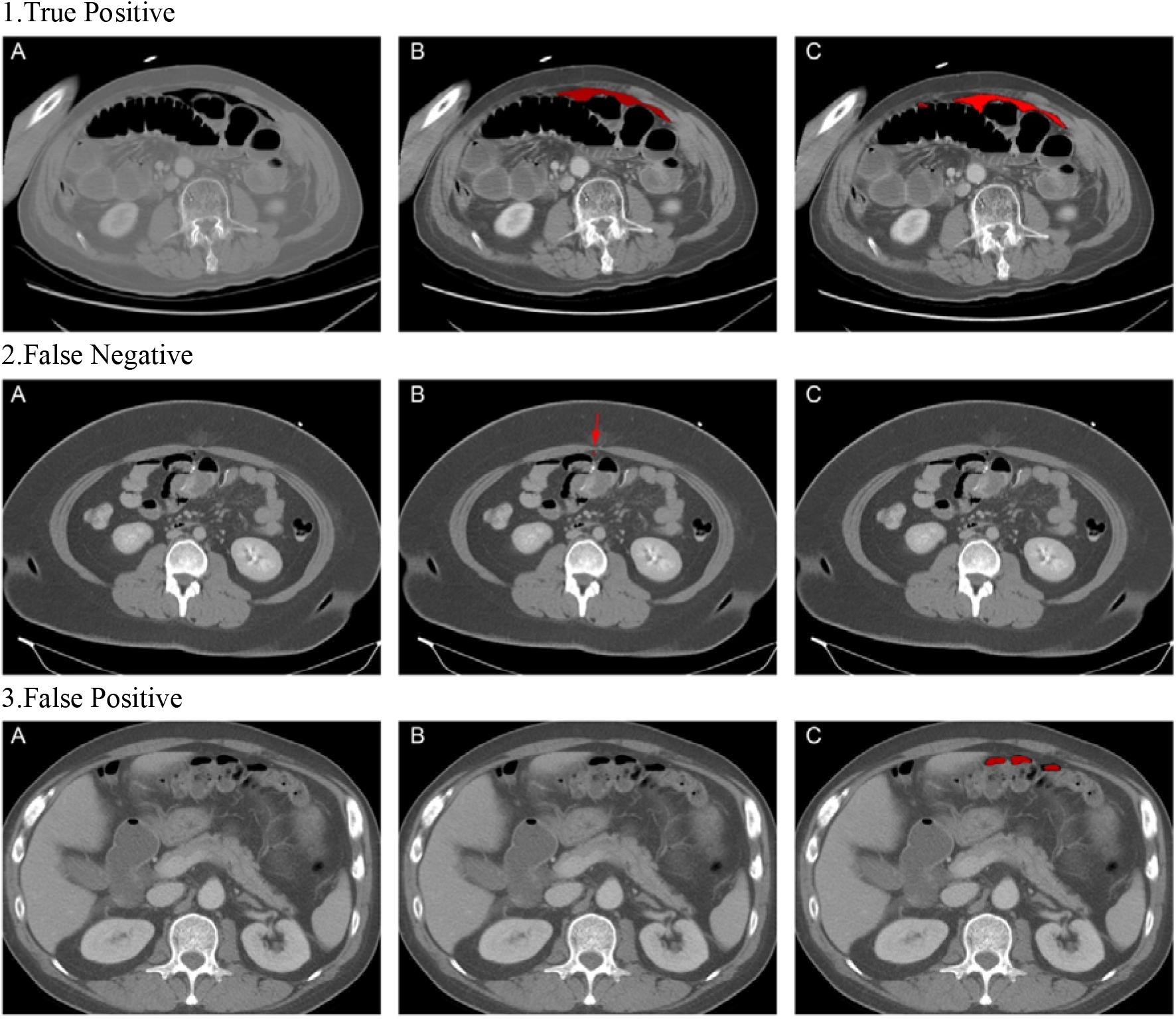
This figure illustrates three distinct outcomes of the model inference in the simulated test set, namely, “True Positive”, “False Negative”, and “False Positive”. For each scenario: ‘A’ represents the original CT scan image, ‘B’ denotes the ground truth labeling, and ‘C’ illustrates the mask generated by the trained segmentation model.

To augment the data, we normalized all CTs to 512×512×z-axis and randomly cubed them to 384×384×z-axis using the ‘albumentations’ library for each image in the training set^26^. The loss function we employed for the model combined Dice loss and Focal loss, each weighted at 50%. This approach aided in addressing class imbalance and enhanced accuracy for hard-to-classify examples^27^. We used an adaptive moment estimation (Adam) optimizer with parameter settings of β1 = 0.9 and β2 = 0.999, and a CosineAnnealingLR scheduler with parameter settings of T_max=8 and eta_min=3 × 10^−6^. The model was trained with the Nvidia RTX A6000 GPU, with minibatches of size 1 and an initial learning rate of 3 × 10^−4^.

### 2.6 Performance Evaluation and Statistical Analysis

The study aimed to evaluate the performance of the PACT-3D model in diagnosing pneumoperitoneum from abdominal CT scans, with continuous variables reported as means and SD, and categorical variables as counts and percentages. The model was trained to minimize loss within the validation dataset, and the optimized model weights were preserved for subsequent inference.

To assess the model’s efficacy, we evaluated its predictive performance on both a simulated test set and a prospective test set. Our primary metrics for evaluation included F1-score, sensitivity, specificity, and PPV, were calculated alongside their 95% confidence intervals. Additionally, we conducted a subgroup analysis to explore how the model’s performance varied across different etiologies such as gastroduodenal, small bowel, large intestine perforations, trauma, and post-operative cases. The modeling pipeline was implemented using Python (3.9) with PyTorch (2.0) and MONAI (1.3.0) as the deep learning framework. Image processing and data analysis were facilitated by Python libraries such as SimpleITK (2.2.1), scikit-image (0.20.0), pandas (2.0.2), and matplotlib (3.7.1), while SPSS was utilized for all subsequent statistical analyses.

## Data Availability

All data produced in the present study are not available for public use

## Notes

### Competing Interest Statement

The authors have declared no competing interest.

### Funding Statement

This study did not receive any funding

### Author Declarations

This study received approval from the Institutional Review Board of Far Eastern Memorial Hospital (FEMH 111086-F).

