## Supplemental Table for "PACT-3D, a Deep Learning Algorithm for Pneumoperitoneum Detection in Abdominal CT Scans"

Supplemental Table 1. Image Acquisition Setting in Different CT Manufacturer

| Manufacturer | Philips | GE | Siemens | Siemens | Siemens | GE |
| --- | --- | --- | --- | --- | --- | --- |
| Scanner model | Brilliance 64 | LightSpeed VCT | Somatom definition | Somatom definition Flash | Somatom definition AS | Revolution Frontier |
| Detector rows | 64 | 64 | 64 | 256 | 64 | 128 |
| Scan mode | Spiral | Helical | Spiral | Spiral | Spiral | Helical |
| kVP | 120 | 120 | 120 | 120 | 120 | 120 |
| Display field of view(cm) | 40-50 | 40-50 | 40-50 | 40-50 | 40-50 | 40-50 |
| Filter type | Body | Body | Body | Body | Body | Body |
| Convolutional Kernel | Standard | Standard | B30f medium smooth | B30f medium smooth | B30f medium smooth | Standard |
| Contrast amount (cc) | <49kg:60cc, 50~69kg:80cc, 70-89kg:100cc, >90kg:120cc | | | | | |
| Contrast injection protocol | Injection speed: 2cc per second | | | | | |

Supplemental Table 2. Performance of Different Sample Ratios in Model Training

| **Positive to Negative Ratio** | **Sensitivity** | **Specificity** | **PPV** | **F1-Score** |
| --- | --- | --- | --- | --- |
|  | value (95% CI) | value (95% CI) | value (95% CI) | value (95% CI) |
| 1: 0 | 0.90 (0.85-0.94) | 0.83 (0.77-0.88) | 0.05 (0.03-0.07) | 0.09 (0.06-0.13) |
| 1: 0.5 | 0.86 (0.81-0.91) | 0.95 (0.91-0.98) | 0.15 (0.12-0.18) | 0.26 (0.28-0.34) |
| 1: 1 | 0.81 (0.75 – 0.86) | 0.99 (0.98-1.0) | 0.41 (0.34-0.48) | 0.54 (0.47-0.61) |
